# Structural mapping of *GABRB3* variants reveals genotype-phenotype correlations

**DOI:** 10.1101/2021.06.04.21256727

**Authors:** Katrine M Johannesen, Sumaiya Iqbal, Milena Guazzi, Nazanin A Mohammadi, Eduardo Pérez-Palma, Elise Schaefer, Anne De Saint Martin, Marie Therese Abiwarde, Amy McTague, Roser Pons, Amelie Piton, Manju A Kurian, Gautam Ambegaonkar, Helen Firth, Alba Sanchis-Juan, Marie Deprez, Katrien Jansen, Liesbeth De Waele, Eva H Briltra, Nienke E Verbeek, Marjan van Kempen, Walid Fazeli, Pasquale Striano, Federico Zara, Gerhard Visser, Hilde M H Braakman, Martin Haeusler, Miriam Elbracht, David Sternman, Ulvi Vaher, Thomas Smol, Johannes R Lemke, Konrad Platzer, Joanna Kennedy, Karl Martin Klein, Ping Yee Billie Au, Kimberly Smyth, Julie Kaplan, Morgan Thomas, Malin K Dewenter, Argirios Dinopoulos, Arthur J Campbell, Dennis Lal, Damien Lederer, Vivian W Y Liao, Philip K Ahring, Rikke S. Møller, Elena Gardella

**Affiliations:** Department of Epilepsy Genetics and Personalized Treatment, the Danish Epilepsy Centre, Dianalund, Denmark; Department of Regional Health Research, University of Southern Denmark, Odense, Denmark; Center for the Development of Therapeutics, Broad Institute of MIT and Harvard, Cambridge, USA; Stanley Center for Psychiatric Research, Broad Institute of MIT and Harvard, Cambridge, USA; Analytic and Translational Genetics Unit, Massachusetts General Hospital, Boston; Department of Medicine, University of Genoa, Genoa, Italy; Department of Clinical neurophysiology, the Danish Epilepsy Centre, Dianalund, Denmark; Universidad del Desarrollo, Centro de Genética y Genómica, Facultad de Medicina Clínica Alemana, Santiago de Chile, Chile; Service de Génétique Médicale, Hôpitaux Universitaires de Strasbourg, Institut de Génétique Médicale d’Alsace, Strasbourg, France; Department of Pediatric Neurology, Strasbourg University Hospital, Strasbourg, France; Molecular Neurosciences, Developmental Neurosciences Programme, UCL Great Ormond Street Institute of Child Health, London, United Kingdom; Department of Neurology, Great Ormond Street Hospital, London, United Kingdom; First Department of Pediatrics, “I Agia Sofia” Children Hospital, National & Kapodistrian University of Athens, Athens, Greece; Laboratoire de diagnostic génétique, Hôpital Civil, CHRU Strasbourg, Strasbourg, France; Department of Paediatric Neurology, Child Development Centre, Addenbrookes Hospital, Cambridge, United Kingdom; Department of Clinical Genetics, Cambridge University Hospitals, Cambridge, United Kingdom; Department of Haematology, NIHR Bioresource, University of Cambridge, United Kingdom; Université Côte d’Azur, CNRS, IPMC, Sophia-Antipolis, France; Department of Pediatric Neurology, University Hospitals Leuven, Belgium; Department of Development and Regeneration, Campus Kulak Kortrijk, Kortrijk, Belgium; Department of Paediatric Neurology, University Hospitals Leuven, Leuven, Belgium; Department of Genetics, University Medical Center Utrecht, Utrecht University, Utrecht, The Netherlands; Department of Pediatric Neurology, University Hospital Bonn, Bonn, Germany; IRCCS Institute “Giannina Gaslini”, Genova, Italy; Department of Neurosciences, Rehabilitation, Ophthalmology, Genetics, Maternal and Child Health, University of Genova, Genova, Italy; Laboratory of Neurogenetics and Neuroscience, IRCCS “G. Gaslini” Institute, Genova, Italy; Stichting Epilepsie Instellingen Nederland (SEIN), Hoofddorp, The Netherlands; Department of Pediatric Neurology, Amalia Children’s Hospital, Radboud University Medical Center, Nijmegen, the Netherlands; Department of Neurology, Academic Center for Epileptology Kempenhaeghe & Maastricht University Medical Center, Heeze, The Netherlands; Division of Neuropediatrics and Social Pediatrics, Dept. of Pediatrics, University Hospital RWTH Aachen, Germany; Institute of Human Genetics, Medical Faculty, RWTH Aachen University, Aachen, Germany; Division of Neurology Lincoln Medical and Mental Health Center, Bronx, NY, United States; Children’s Clinic of Tartu University Hospital, Tartu, Estonia; ERN EpiCARE, Tartu, Estonia; Institut de Genetique Medicale, CHRU Lille, Universite de Lille, Lille, France; Institute of Human Genetics, University of Leipzig Medical Center, Leipzig, Germany; Clinical Genetics Department, University Hospitals Bristol NHS Foundation Trust, St Michael’s Hospital, Bristol, United Kingdom; Departments of Clinical Neurosciences, Medical Genetics and Community Health Sciences, Hotchkiss Brain Institute & Alberta Children’s Hospital Research Institute, Cumming School of Medicine, University of Calgary, Calgary, Canada; Epilepsy Center Frankfurt Rhine-Main, Department of Neurology, Center of Neurology and Neurosurgery, University Hospital, Goethe-University Frankfurt, Germany and Center for Personalized Translational Epilepsy Research (CePTER), Goethe University Frankfurt, Germany; Department of Medical Genetics, Alberta Children’s Hospital Research Institute, Cumming School of Medicine, University of Calgary, Calgary, Canada; Department of Pediatrics, Cumming School of Medicine, University of Calgary, Calgary, Canada; Division of Medical Genetics, Nemours A.I. duPont Hospital for Children, Wilmington, Delaware, USA; Precision Medicine/Genetic Testing Stewardship Program, Nemours A.I. duPont Hospital for Children, Wilmington, Delaware, USA; Institute of Human Genetics, Universitätsmedizin, Johannes Gutenberg-University, Mainz Institut für Humangenetik, Mainz, Germany; Attiko University Hospital, University of Athens, 3rd Dept of Pediatrics, Haidari, Greece; Genomic Medicine Institute, Lerner Research Institute, Cleveland Clinic, Cleveland, OH, USA; Epilepsy Center, Neurological Institute, Cleveland Clinic, Cleveland, OH, USA; Cologne Center for Genomics (CCG), University of Cologne, Cologne, Germany; Centre de Génétique Humaine, Institut de Pathologie et de Génétique, Charleroi, Gosselies, Belgium; Brain and Mind Centre, School of Pharmacy, Faculty of Medicine and Health, The University of Sydney, Sydney, New South Wales, Australia

**Keywords:** Epilepsy, Genetics, GABA, GABRB3, mapping

## Abstract

**Purpose:** Pathogenic variants in *GABRB3* have been associated with a spectrum of phenotypes from severe developmental disorders and epileptic encephalopathies to milder epilepsy syndromes and mild intellectual disability. In the present study, we analyzed a large cohort of individuals with *GABRB3* variants to deepen the phenotypic understanding and investigate genotype-phenotype correlations.

**Methods:** Through an international collaboration, we analyzed electro-clinical data of unpublished individuals with variants in *GABRB3* and we reviewed previously published cases. All missense variants were mapped onto the 3D structure of the *GABRB3* subunit and clinical phenotypes associated with the different key structural domains were investigated.

**Results:** We characterize 71 individuals with *GABRB3* variants, including 22 novel subjects, expressing a wide spectrum of phenotypes. Interestingly, phenotypes correlated with structural locations of the variants. Generalized epilepsy, with a median age at onset of 10.5 months, and mild-to-moderate intellectual disability were associated with variants in the extracellular domain. Focal epilepsy with early onset (median: 2.75 months of age) and severe intellectual disability were associated with variants in the pore-lining helical transmembrane domain.

**Conclusion:** These genotype/phenotype correlations will aid the genetic counseling and treatment of individuals affected by *GABRB3*-related disorders. Future studies may reveal whether functional differences underlie the phenotypic differences.

**Key points:**

- Pathogenic variants in *GABRB3* cause a wide range of phenotypes
- Missense variants in the ECD have generalized epilepsy with later onset and non-severe ID
- Missense variants in the TMD have focal epilepsy with early onset and severe ID
- Behavioral issues are common features of *GABRB3* disease
- Precision medicine approaches for *GABRB3* disease is limited

## Introduction

The *GABRB3* gene encodes the β3 subunit of the ligand-gated γ-amino-butyric-acid type A receptor (GABA_A_R). The β3 subunit is one of the most abundant subunits in the human central nervous system, particularly during early stages of life, and therefore has a crucial role in neurodevelopment during the embryonic state^1^. Being abundantly expressed, the β3 subunit also plays an important part in regulating the number of receptors in the synapse during inhibitory synaptic plasticity and is critical for the pentameric receptor assembly^2^.

It has recently been reported that individuals with pathogenic *GABRB3* variants present a broad phenotypic spectrum from severe developmental and epileptic encephalopathy (*de novo* variants) to milder epilepsy syndromes including generalized epilepsy with febrile seizures+ (GEFS+) (sometimes familial) and childhood absence epilepsy^3^. The current consensus dictates that pathogenic variants in *GABRB3* cause loss of GABA_A_R activity. When GABA_A_Rs cannot effectively open, neuronal hyperactivity occurs, which ultimately increase seizure susceptibility and lead to various neurological and behavioral abnormalities^4-6^. Several mechanisms have been suggested whereby variants in the β3 subunit cause structural changes in the protein resulting in receptor malfunction or irregular expression levels in synapses^2, 7, 8^. Given the observed broad phenotypic spectrum in individuals with pathogenic variants in *GABRB3*, it is assumed that one or combinations of several of these mechanisms come into play to cause the phenotypic differences^2, 4, 5, 7^.

Over the years, extensive analysis of the three-dimensional (3D) structure of α1β3γ2 GABA_A_R has revealed important structure-function relationships, most of which was corroborated by recent CryoEM structures of the pentameric receptor complex^9^. Each subunit has an extracellular domain followed by helical M1 – M4 regions forming the transmembrane domain, with M2 segments lining the channel pore. GABA_A_R channels open upon binding of GABA molecules to specific binding sites at the interfaces between β3/α1 subunits. This causes an influx of chloride ions into the cell hence decrease neuronal excitability. Given that various structural regions in a subunit play specific roles in receptor activation, it can be speculated that pathogenic variants are predominantly located in these regions. Therefore, analyzing variants in the context of the 3D α1β3γ2 GABA_A_R structure will provide insight into their differential phenotypic outcome in *GABRB3*-related disorders.

To provide a comprehensive review of all available individuals with pathogenic *GABRB3* variants, we here collate the knowledge of all known individuals and extend this with a new cohort of 22 individuals. Furthermore, by correlating the 3D structural locations of the variants with clinical features, we investigated whether certain structural regions were more likely to be associated with more severe phenotypes.

## Materials and methods

Through an international collaboration including epilepsy centers in Europe and North America, we collected 22 unpublished individuals with *GABRB3* variants. The ACMG/AMP guidelines were used to classify variant pathogenicity^10^. The *GABRB3* transcript number NM_000814.5 was used for coding variant nomenclature.

Clinical information was collected by face-to-face interviews with the affected individuals and their caregivers and from clinical charts. The referring clinicians collected all data by a structured phenotype table, which included cognitive and motor milestones, details about epilepsy and EEG, as well as treatment response. The epilepsy syndromes and seizure types were classified according to the guidelines of the International League Against Epilepsy^11^. Epilepsy types were classified as generalized, focal or unknown, based on clinical grounds, supported by EEG findings (epilepsydiagnosis.org)^12^.

EEG reports at seizure onset and at several points of follow up were obtained for all novel individuals. A single epileptologist (EG) with EEG expertise reviewed raw EEG data of 13 affected individuals (including long-term monitoring video-EEGs) for background activity, interictal epileptiform abnormalities, ictal EEG discharges and clinical manifestations.

### Review of the literature

In parallel, we reviewed all available data on previously published *GABRB3* individuals. The literature search was performed using Pubmed and Scopus, key words included *GABRB3*, epilepsy, autism spectrum disorder and psychiatric features. Papers in non-English language were excluded. Last search date was March 1^st^ 2021.

### Structure of GABA_A_R α1β3γ2

Three-dimensional (3D) crystallographic structure for human GABA_A_R β3 in a homopentameric form (PDB ID: 4COF^13^), resolution = 2.97Å, residue position coverage = 26 – 332 and 447 – 473) in complex with an agonist (benzamidine) was obtained from the Protein Data Bank^13^. Additionally, we performed our analysis on the biological assembly of GABA_A_R (PDB ID: 6I53^9^, a Cryo-EM structure of the human α1β3γ2 GABA_A_ receptor in a lipid bilayer, resolution = 3.20Å, residue position coverage = 26 – 473). Mapping of gnomAD v2.1.1 missense variants and *GABRB3* variants on the structure was performed with PyMOL.

## Results

We collected electro-clinical data on a total of 71 affected individuals with pathogenic or likely pathogenic *GABRB3* variants^3, 8, 14-30^, including 22 previously unpublished individuals. Median age at follow-up was six years (range 10 months to 46 years). The main electro-clinical and genetic features are summarized in Table E1. Complete information was not available in all individuals; denominators indicate the individuals in whom information on the clinical feature addressed was available.

### Epilepsy

99% (70/71) of all individuals suffered from epilepsy. 45% (19/42) of the individuals suffered from generalized epilepsy (GE) and 55% (23/42) had focal epilepsy (FE). In 29 individuals the epilepsy type was unknown. Epilepsy syndromes varied from individual to individual (Fig. 1A). The median age of epilepsy onset was 10.5 months (range: 4 months – 5.2 years) in individuals with GE and 2.75 months (range: day 3 - 17 months) in individuals with FE (Fig.1B).

**Figure 1.**
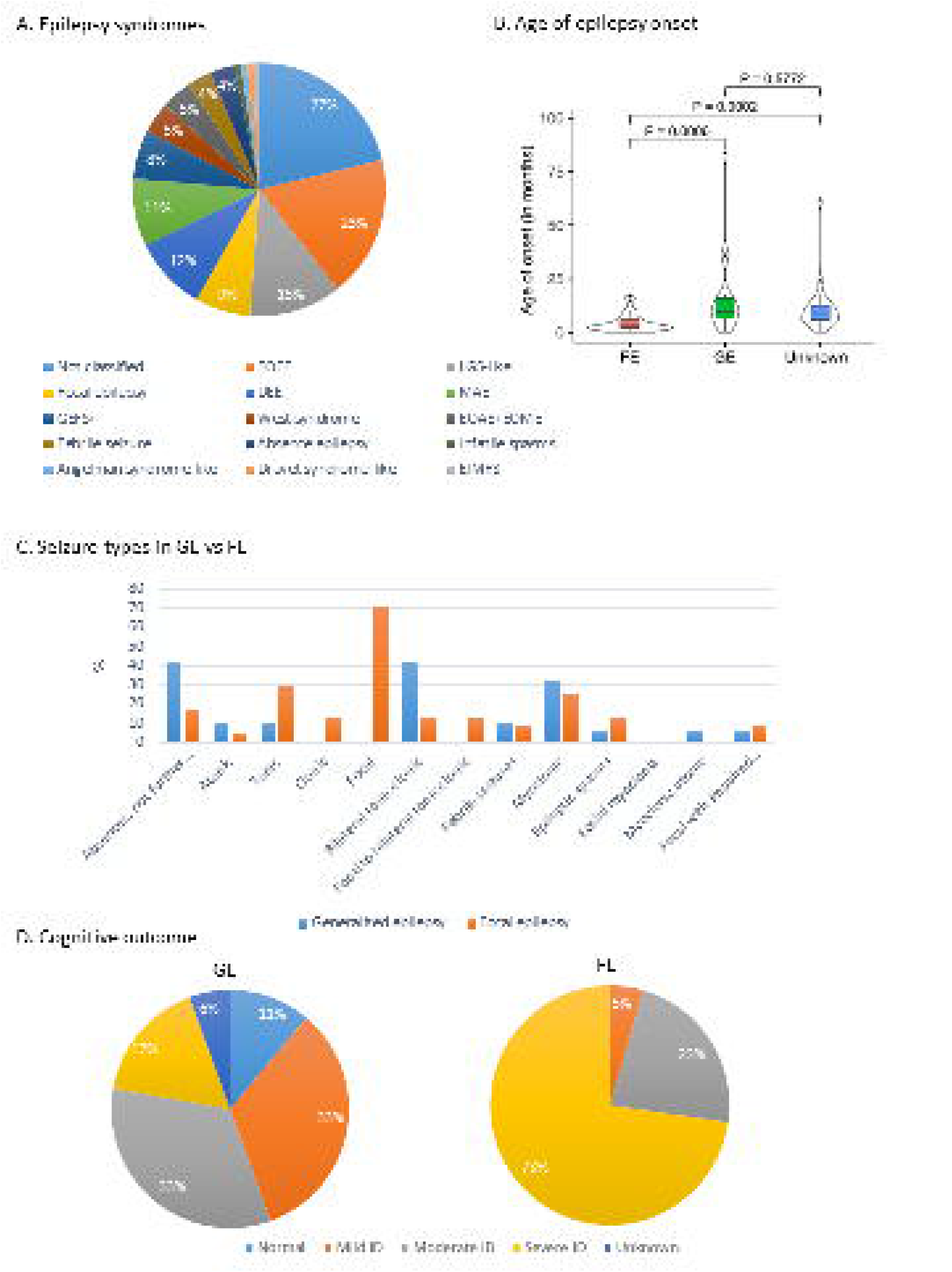
A. Epilepsy syndromes in the *GABRB3* cohort B. Seizure onset in individuals with generalized vs. focal epilepsy C. Seizure types in individuals with generalized vs. focal epilepsy D. Cognition in individuals with generalized epilepsy and with focal epilepsy

Each individual presented with up to five different seizure types, including focal seizures (29%, 20/70), bilateral tonic-clonic seizures (29%, 20/70) and myoclonic seizures (27%, 19/70) (Fig. 1C). Status epilepticus was reported in 13% (9/70) individuals. Fever sensitivity was found in 29% (19/70) individuals.

Several anti-seizure medications (ASMs) in various combinations were tested in each individual. At latest follow up, 51% (26/51) of the individuals had ongoing daily to weekly seizures and 41% (21/51) were seizure free or had rare seizures. 37% (7/19) of the individuals with GE achieved seizure freedom, either with valproate (VPA) polytherapy (n=3), or with lamotrigine and clobazam (CLB), barbiturates, clonazepam (CZP) or steroids (n=1 each). 17% (4/23) of individuals with FE achieved seizure freedom with VPA, CZP or stiripentol, levetiracetam (LEV), alone or in combination with VPA, or CLB and vigabatrin (VGB) (n=1 each).

### EEG

The interictal EEG at epilepsy onset was normal in 32% (7/22) of individuals. In the remaining individuals, the EEG showed background slowing (14%, 3/22) and irregular generalized spike and waves (14%, 3/22), multifocal (14%, 3/22) or focal spike/sharp-and-slow waves (14%, 3/22), hypsarrhythmia (14%, 3/22), or a burst suppression pattern (9%, 2/22).

At follow-up, the EEG deteriorated in most of individuals, remaining normal in only 9% (2/22) of individuals, and showed background slowing (18%, 4/22), and focal spikes (36%, 8/22), or multifocal/generalized spike and waves (27%, 6/22), burst suppression (5%, 1/22), hypsarrhythmia (5%, 1/22), migrating focal seizures of infancy (9%, 2/22).

In five individuals, ictal EEG recordings were available, including focal seizures (Fig.2A) in three individuals with focal epilepsy and tonic seizures during sleep with ictal generalized fast activity (Fig.2B) in two individuals with generalized epilepsy.

**Figure 2.**
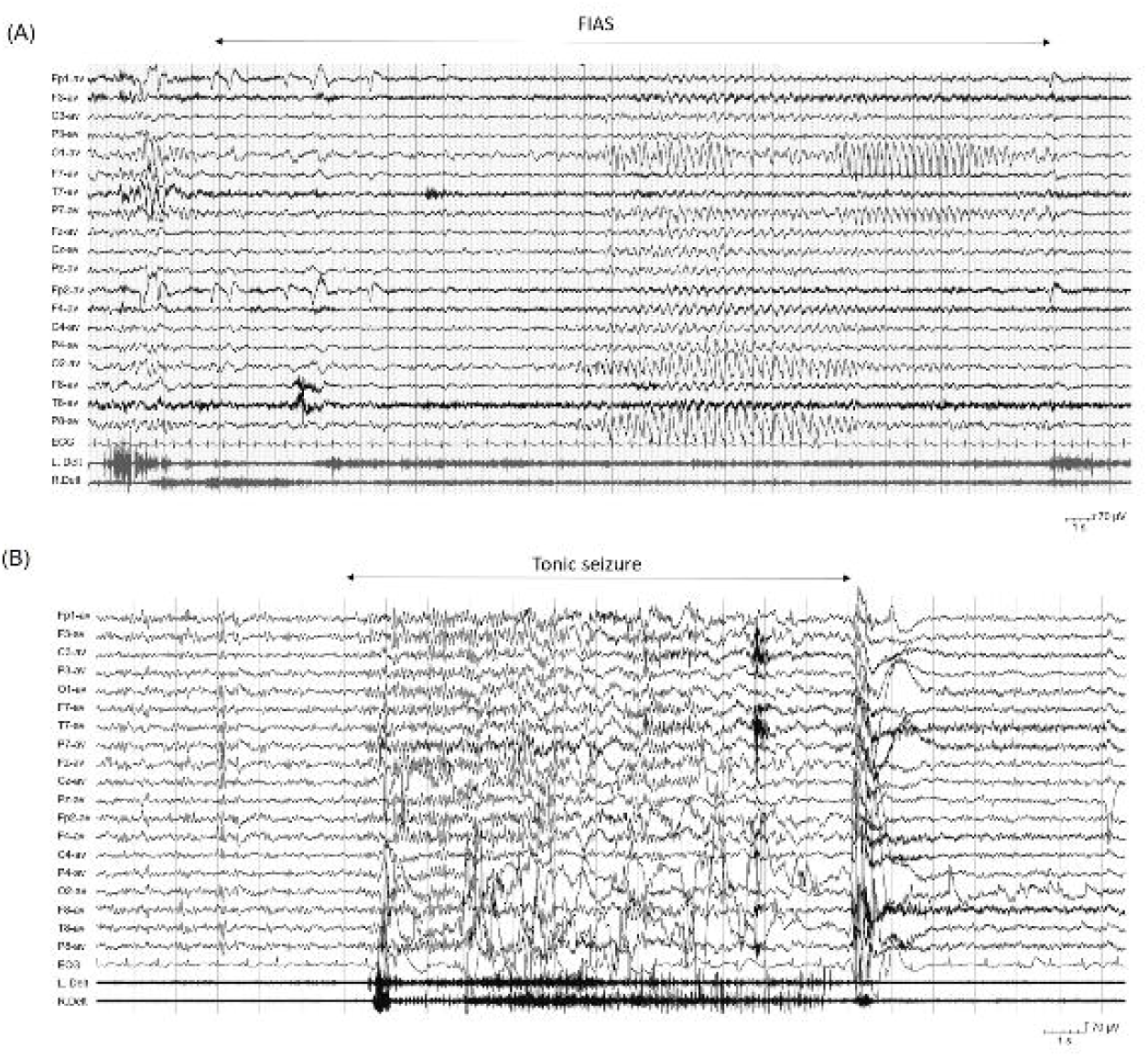
Ictal EEG recordings in individuals with focal epilepsy (FE) or with generalized epilepsy (GE) related to *GABRB3* pathogenic variants. (A) Individual #9 (FE): focal seizure during wakefulness with “staring” and impaired awareness (FIAS). The EEG shows an arrest of the background activity, followed by low amplitude rhythmic activity in the posterior regions, evolving to high amplitude 6 Hz activity with intermixed small spikes, in the occipital-posttemporal regions bilaterally. (B) Individual #8 (GE): tonic seizure during sleep. The EMG traces to the bottom show that the seizure starts with a spasm, followed by sustained tonic activation, with a superimposed vibratory component close to the seizures’ end. The EEG correlate is a diffuse 10 Hz rhythmic activity with frontal predominance.

### Cognition, behavior and additional features

At latest follow up, 39% (28/71) of the individuals had severe ID, 32% (23/71) had moderate ID, while 17% (12/71) had mild ID and 3% (2/71) had normal cognition. Cognitive stagnation or regression after epilepsy onset was described in 25% (18/71) of the individuals.

Individuals with FE had a higher prevalence of severe ID (75%, 18/24), compared to those individuals with GE (16%, 3/19) (Fig.1D).

Autism spectrum disorder was diagnosed in 5% (4/71) of the individuals (one GE, four unknown) and autistic features were seen in 15% (11/71) (one GE, four FE, six unknown). Behavioral issues were present in 11% (8/71) of the individuals (four GE, one FE), including aggression, impulsivity, anxious behaviors or oppositional defiant disorders. Attention Deficit Hyperactivity Disorder (ADHD) was reported in 10% (7/71) of the individuals, and 8% (6/71) had stereotypic behavior. Neurological disturbances were observed in 42 individuals (9 GE, 22 FE), consisting of axial hypotonia (40%, 17/42), ataxia (17%, 7/42), dystonia/dyskinesia (10%, 4/42), delayed speech and language development (55%, 23/42), microcephaly (12%, 5/42), and central visual impairment (14%, 6/42). Sleep disturbance were reported in 14 % (6/42) of individuals.

Dysmorphic features were reported in a subset of individuals (11%, 8/71) and included a mild prominence of the forehead, tented mouth appearance and high-arched palate. One individual had cleft-palate^30^.

### Affected family members

We also collected information about additional family members carrying pathogenic *GABRB3* variants. All harbored protein-truncating variants (PTVs). 54% (13/24) were asymptomatic and 46% (11/24) had epilepsy. All but one family member had normal intellect (data available for 73% (8/11) in the epilepsy group, and for 92% (11/12) in the no-epilepsy group); one was deceased and thus unavailable for follow-up. One family member had cognitive regression after seizure onset. No data about neurological or behavioral issues were available.

### Genetic landscape

A total of 53 different variants was observed in the 71 patients. Eleven of them carried ten different PTVs, in the form of seven stop-gained, two frameshift variants and one in-frame duplication (duplication of one amino acid), while 59 patients carried 42 different missense variants (single base substitution leading to a single amino acid change in the protein). Only one patient had a small indel variant leading to a single amino acid change in the protein. *GABRB3* has a high pLI score for PTVs (ratio of expected versus observed variants) of 0.95, and only very few PTVs in gnomAD, indicating that the gene is intolerant for PTVs.

In the rest of the paper, all single amino acid changing variants are referred to as missense variants. 76% (54/71) of the individuals had *de novo* variants, 17% (12/71) had variants inherited from a parent, either affected or unaffected, while for five individuals, segregation was unknown. Thirteen variants were recurrent, displaying homogenous phenotypes for identical variants.

Missense variants were primarily located in the N-terminal extracellular domain (ECD; 23 variants in 35 individuals) and in the transmembrane domain (TMD; 20 variants in 25 individuals) (Fig. 3). A few variants were located in the large intracellular loop (ICD) between M3 and M4 (two variants in two patients). In the TMD, variants were mainly clustered in two regions: three helical segments (M1, M2, M3; 16 variants) and a key structural coupling region (small M2 – M3 loop; three variants). All inherited variants were located in the ECD (11 individuals) except one in the M4 (one individual) helical segment of the TMD (Fig.3A).

**Figure 3.**
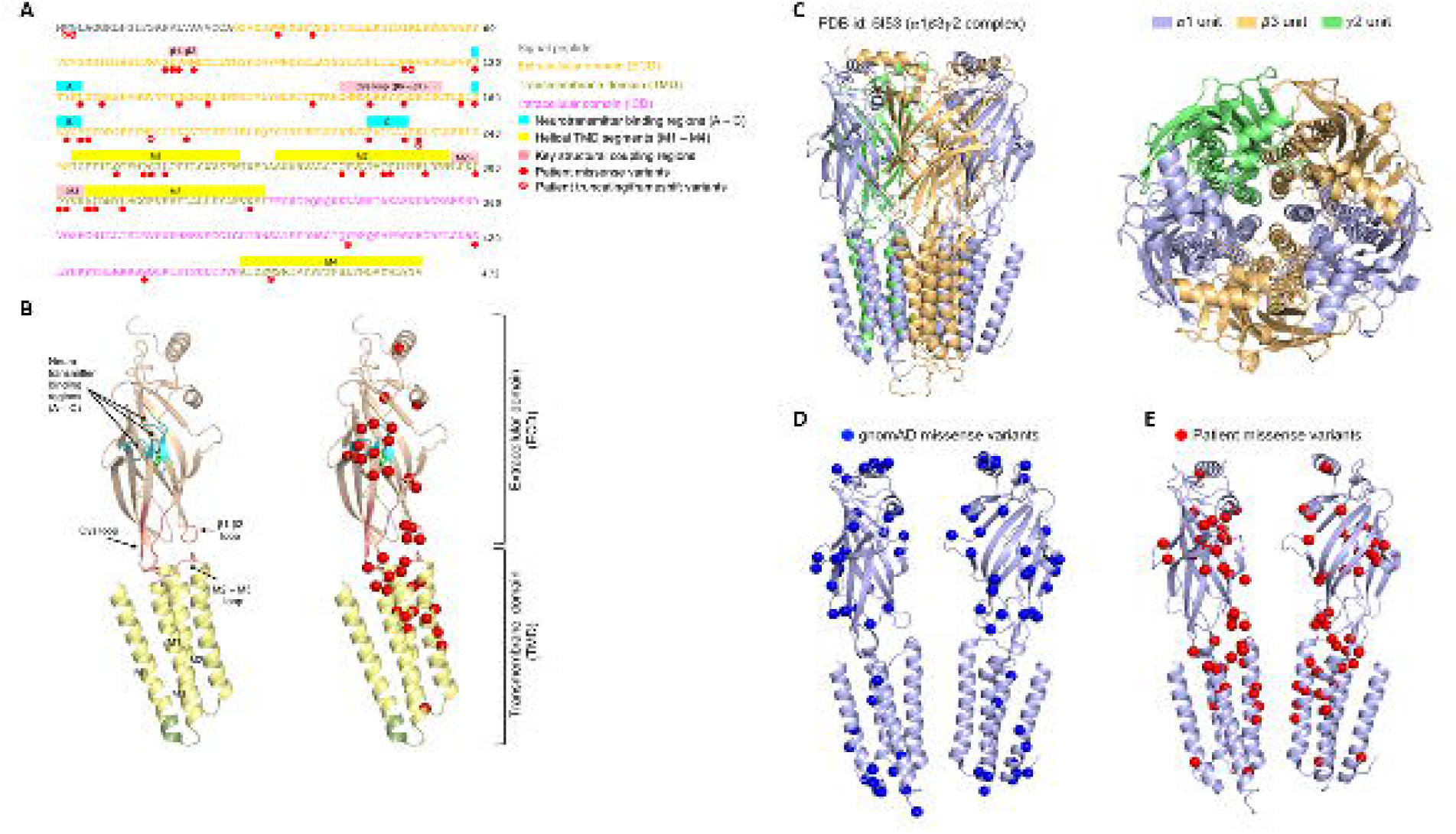
(A) Protein sequence encoded by *GABRB3*, annotated with protein domains, neurotransmitter binding regions and helical segments, structural coupling regions and individual variants’ positions. (B) GABA_A_R β3 structure (single chain, PDB ID: 4COF), annotated with protein domains, neurotransmitter binding regions and helical segments, structural coupling regions and individual missense variants’ positions (red spheres). (C) Side (left) and top (right) view of the Cryo-EM structure of the human synaptic α1β3γ2 GABA_A_R complex (PDB id: 6I53). Two α1 chains (encoded by **GABRA1**), two β3 chains (encoded by *GABRB3*) and one γ2 chain (encoded by **GABRG2**) are shown in light orange, purple and green, respectively. (D) *GABRB3* missense variants observed in the general population (gnomAD database) mapped onto β3 chains (blue spheres). (E) *GABRB3* missense variants observed in individuals (our cohort, table E1) mapped onto β3 chains (red spheres).

### Individual and population missense variants and structural locations

We mapped the location of the missense variants from our cohort and population missense variants from the gnomAD database onto the GABA_A_R β3 sequence (UniProtKB - P28472) and structure (PDB ID: 4COF, an X-ray structure, Fig. 3B, and PDB ID: 6I53, a Cryo-EM structure, Fig. 4A). In the general population, 110 variants were observed which are located in 91 amino acid positions according to the gnomAD database^31^. 43% (44 out of 110) of these general population variants were mappable on the transmembrane structure (Fig. 3D), while the rest were located in the unstructured regions of the extracellular domain (ECD) and in the M3 – M4 region of the intercellular domain (ICD).

**Figure 4.**
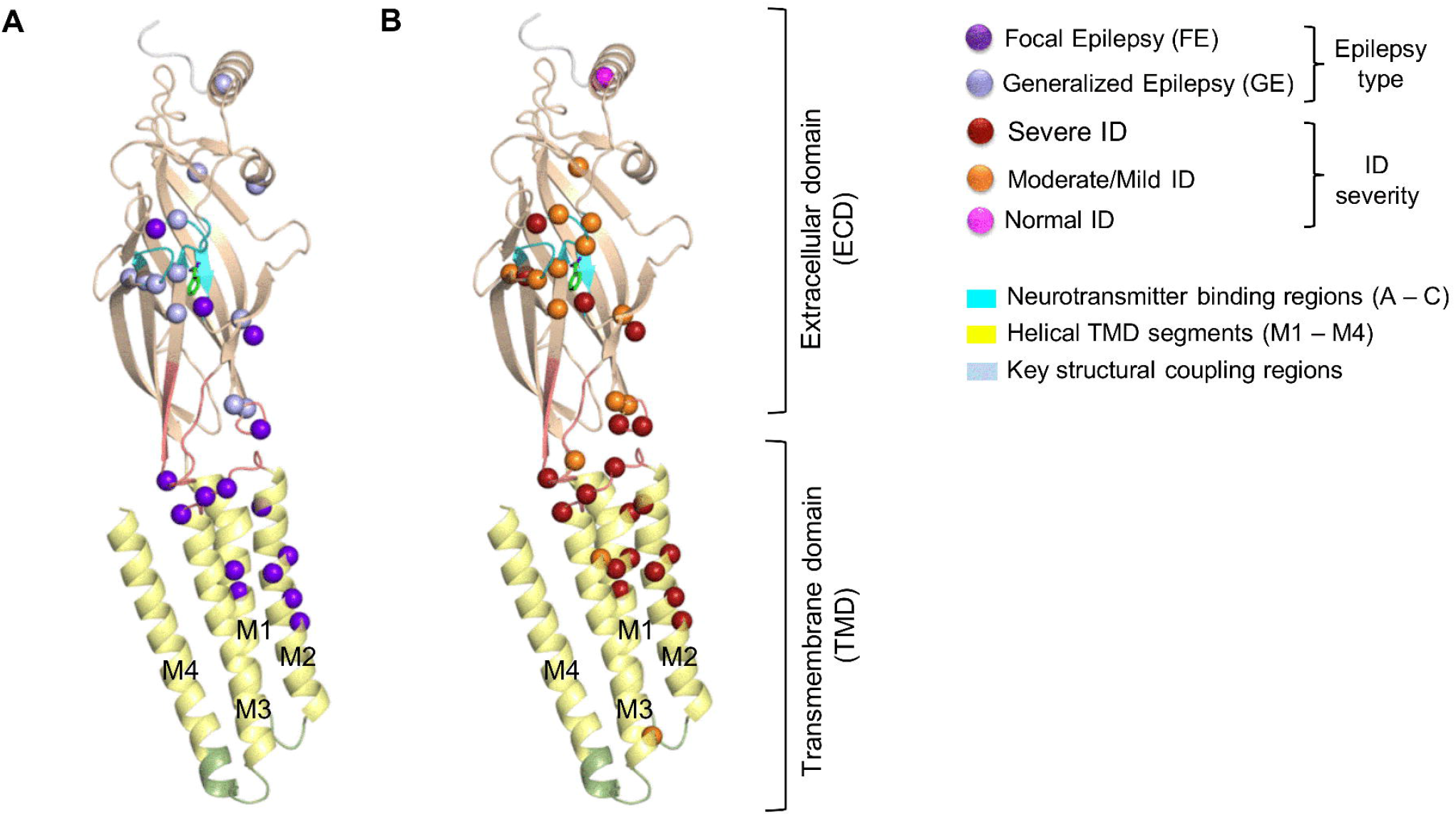
Investigation of individual missense variant positions by different phenotypes on the GABA_A_R β3 structure (PDB ID: 4COF, single chain). (A) Protein residue positions affected by missense variants in individuals with different epilepsy types: focal epilepsy and generalized epilepsy are indicated by dark purple and light purple, respectively. (B) Protein residue positions affected by missense variants in individuals with different severity of ID: severe ID, moderate/mild ID and normal ID are indicated by dark red, orange and magenta spheres, respectively.

In contrast, 95% (41/43) disease-associated missense variants, substituting 38 amino acid residues, were mappable to the structure (Fig.3B and Fig. 3E). Only two variants were located in the unstructured M3 – M4 region of the ICD, which were not available in the crystal structure or in the Cryo-EM structure (Table E1); these are *de novo* variants, p.(Ser420Ile) and p.(Ser433Leu).

Two amino acid positions were mutated in both affected individuals and in the general population (Asn110 and Arg142).Overall, it is notable that the disease-associated missense variants were located in distinct spatial regions of the structure (hotspots) compared to the general population variants (Fig. D-E); preferentially on or in a spatial proximity to: (1) the neurotransmitter binding regions A, B, and C in the ECD, which are essential for the neurotransmitter recognition and binding (Fig. 3B); (2) the β1 – β2, Cys, and M2 – M3 loops, which are key structural regions for coupling and signaling between the ECD and TMD; and (3) the channel forming helical segments M1, M2 and M3 (Fig. 3B).

### Are individual phenotypes correlated with the structural location of the variant?

Given the clustering of variants in key regions of the *GABRB3* protein (Fig. 3), it is likely that the structural position of a variant is correlated with the individual phenotype. To investigate this, we grouped the individuals with a missense variant (n = 64) based on their ID severity and epilepsy type (Table 1) and mapped the affected amino acids on the structure (Fig. 4). All 18 individuals with GE in our cohort had missense variants located in the ECD of the protein (Fig. 4A), except p.(Ser433Leu) which is located in the intracellular loop between M3 and M4 helical regions. Conversely, FE associated missense variants are located in both the ECD and TMD. All individuals with a variant in the M2 helical segment of TMD had FE (Fig. 4A) and severe ID (Fig. 4B). Hence, it appears that at least certain clinical phenotypes are indeed associated with the structural location of the missense variant.

**Table 1.** Clinical characteristics of the *GABRB3* cohort

|  | Missense variants |  | Truncating variants |
| --- | --- | --- | --- |
|  | ECD | TMD* |  |
| Number of individuals / variants | 35 / 23 | 24 / 20 | 10 / 9 |
| Epilepsy onset (range) | 9 months<br>(1.5 m – 7 y) | 4 months<br>(birth – 12 y) | 1 year<br>(birth – 5 y, 2 m) |
| Seizure types (top 3) | Myoclonic (31%)<br>Atonic (29%)<br>Bilateral TC (29%) | Focal (50%)<br>Tonic (33%)<br>Myoclonic (21%) | Absences (40%)<br>Bilateral TC (20%)<br>Myoclonic (10%) |
| Epilepsy type (number of individuals) | 17 GE / 6 FE | 1 GE / 15 FE | 2 GE / 1 FE |
| Fever sensitivity | 29% | 13% | 50% |
| Seizure freedom (mean age at offset) | 34% (1.5 y) | 17% (11.5 y) | 40% (5 y) |
| Most effective ASMs | VPA in combinations | None specific | VPA in combinations |
| Cognition | Normal 3%<br>Mild ID 30%<br>Moderate ID 39%<br>Severe ID 27% | Normal 0%<br>Mild ID 10%<br>Moderate ID 14%<br>Severe ID 76% | Normal 10%<br>Mild ID 0%<br>Moderate ID 60%<br>Severe ID 30% |
| Neurological deficits | Language delay (63%)<br>Hypotonia (26%)<br>Ataxia (16%) | Hypotonia (52%)<br>Language delay (29%)<br>Ataxia (19%)<br>Visual impairment (19%) | Language delay (40%) |
| Additional features | ADHD (50%)<br>Stereotypies (33%)<br>Autistic features (23%) | Autistic features (13%)<br>Aggression (8%)<br>Microcephaly (13%)<br>Digestive issues (13%) | Autistic features (40%)<br>Aggression (20%)<br>Stereotypies (20%) |
| Recurrent variants | p.(Asp120Asn), p.(Leu124Phe),<br>p.(Lys127Arg), p.(Thr157Met),<br>p.(Thr185Ile), p.(Arg232Gln) | p.(Ser254Phe), p.(Leu256Gln),<br>p.(Thr281Ala), p.(Pro301Leu),<br>p.(Tyr302Cys) | p.(Trp2*) |

\*Including M2-M3 loop
Abbreviations: ADHD: Attention deficit hyperactivity disorder, ASMs anti-seizure medications, ECD: extracellular domain, FE: focal epilepsy, GE: generalized epilepsy, TC: tonic-clonic seizures, TSM: transmembrane domain, m: months, y: years

### Genotype-phenotype correlations and protein position of missense variants

In the following, we analyzed the individual phenotypes of missense variants (65 individuals) by separating them into three groups based on key structural locations ECD, TMD and ICD (Table 1).

### In the ECD group

All individuals (n=35) suffered from epilepsy with a median age at epilepsy onset of 8.5 months (range 1.5 months to seven years). Half (49%, 17/35) had GE, with fever sensitivity in 29% (10/35) of cases. Seizure freedom was achieved in 34% (12/35) of individuals at a median age of 1.5 years, after trying 1-3 ASMs. The most effective medications included valproate (VPA) (14% (5/35) of individuals) in combination with either topiramate (TPM), levetiracetam (LEV), zonisamide (ZNS), or steroids.

Cognition before epilepsy onset was normal in 71% (12/17) of subjects, while 24% (4/17) had mild to moderate ID; cognition was unknown in 18 subjects. At latest follow up, cognition deteriorated; 3% (1/33) had normal cognition, 70% (23/33) had mild ID to moderate ID and 27% (9/33) severe ID. In two individuals cognition was unknown.

Behavioral issues were common (34%, 12/35) including ADHD (6/12), stereotypic behavior (4/12) and aggression (2/12). Autism or autistic features were reported in 23% (8/35).

Neurological deficits were reported in 54% (19/35): language delay was the most common issue (63%, 12/19), followed by hypotonia (26%, 5/19) and ataxia (16%, 3/19). One individual, carrying a p.(Tyr182Phe) variant, died at 35 months of SUDEP^19^.

### In the TMD group

15 individuals (63%, 15/24) had a FE and one had a GE. The median age at onset was four months (range first day of life to five years). Fever sensitivity was not prominent (13%, 3/24). Seizure freedom was achieved in 17% (4/24) at a median age of 12 months, both with mono- and poly-therapy with LEV, lamotrigine (LTG), phenobarbital (PB), stiripentol (STP), vigabatrin (VGB) and VPA, no ASM was preferred over others.

Cognition at onset was known only for four individuals, being normal in two, while two had mild ID. All individuals were intellectually disabled at latest follow up: 10% (2/21) had mild ID, 14% (3/21) had moderate ID and 76% (16/21) had severe ID. The level of cognition was unknown in three individuals. A few individuals had autistic features (n=3) or behavioral issues (n=2). Neurological deficits were reported in 88% (21/24), mainly consisting of hypotonia (11/21), language delay (6/21), ataxia (4/21), had visual impairment (4/21). Two individuals were deceased: one (p.(Thr288Asn)), died at 18 months in the context of severe neurological deterioration^8^ and one (p.(Leu284Arg)) died at 54 months of unknown causes^20^.

### In the ICD group

Only two individuals carried missense variants in the ICD, not permitting any comparison with the other subgroups.

### Individuals with truncating variants

All individuals (n=10) had epilepsy with a median age at onset of one year, and with prominent fever sensitivity (50%). Seizure freedom was achieved in 40% (4/10), with VPA poly-therapy in two. Cognition before seizure onset was normal in half of the individuals (2/4). At latest follow up, most individuals had moderate (n=6) or severe (n=3) ID. Language delay was the only neurological deficit reported (n=4). Autistic features (n=4) and behavioral issues (n=4) were prominent.

## Discussion

In this study, we analyzed the phenotypic and genetic features of a large cohort of 71 individuals with a *GABRB3*-related disorder, including 22 previously unpublished individuals. We delineated a spectrum of developmental diseases associated with *GABRB3* variants, including various epilepsy phenotypes. Furthermore we found a possible correlation between the location of the variants within the protein structure and the clinical features, including age of epilepsy onset, epilepsy type and degree of ID.

### Clinical and genetic landscape

Like previous studies, when we analyzed the whole cohort, we were unable to delineate a unique *GABRB3* phenotype. We found that phenotypes were distributed across a large clinical spectrum, encompassing a wide variety of epilepsy syndromes both generalized and focal, as well as different degrees of intellectual disability.

Interestingly, by clustering affected individuals based on location of their variants within the protein, we observed more homogeneous clinical subgroups with different phenotypic features and severity. Individuals with variants located in the ECD primarily had GE with onset of myoclonic, atonic or absence seizures at a median age of 10.5 months. Furthermore, 28% of these individuals were fever sensitive. Behavioral issues, including ADHD, and autistic features were reported in half of the individuals (52.8%). The clinical features correlated with the findings in the *GABRB3*^+/D120N^ (extracellular variant) mouse that displays atypical absence seizures, myoclonic seizures, tonic seizures and bilateral TCs, as well as rare atonic seizures and epileptic spasms in the *GABRB3*^+/D120N^ pups^5^. Behavioral issues have also been described for the *GABRB3*^+/D120N^ mouse that have learning and memory deficits, hyperactivity, reduced socialization and anxiety that all increase with age^5^. The few individuals with protein truncating variants displayed a phenotype resembling that found in individuals with variants located in ECD which included generalized seizures, moderate ID and relatively common behavioral issues or autistic features. Interestingly, the majority of the PTVs were inherited from presumed unaffected parents, suggesting reduced penetrance as previously discussed^3^. In contrast, individuals with variants in the TMD displayed a more severe phenotype with early onset FE, often refractory to ASMs and they were usually not fever sensitive. The majority of these individuals had severe ID and prominent neurological deficits, while behavioral issue and autistic features were rarely described.

### Clinical phenotypes in key structural regions

When analyzed in a structural context, it was clear that variants were clustered in three regions of the β3 subunit that are key to its function.

#### GABA binding region

A hotspot of variants was identified in the neurotransmitter binding pocket formed by neurotransmitter binding regions A, B and C^13^. Neurotransmitter (GABA) binding activates the GABA_A_R channel and therefore variants in this pocket should have considerable impact on the activation and/or deactivation of the channel. In total, seven variants (11 individuals) were observed in this pocket in our cohort. Six of these variants led to moderate to mild ID with or without GE. Interestingly, this implies that individuals with variants in the GABA binding pocket are less severely affected relative to those with variants in other structural motifs. One notable exception to this is the p.(Leu124Phe) variant, which was found in three individuals with severe ID and FE, suggesting that this specific location is somehow crucial for the function of the protein.

#### Coupling region

Another important region of the GABA_A_R β3 subunit is the ECD–TMD interface. For a neurotransmitter binding event to transmit to the channel gate, the signal must be transduced across the ECD–TMD interface. Given the importance of this interface for inter-subunit structural coupling and signal transmission, variants in this interface are likely to cause attenuation of the signaling events and thus leading to disease. In our cohort, 10 variants (in 14 individuals) in the ECD–TMD interface showed different phenotypes. This suggests that even if these structural motifs are important for neurotransmitter binding, not all variants in these locations will lead to a severe phenotype.

#### Transmembrane domains

All nine individuals with variants in the M2 had FE and either severe ID (8/9) or moderate ID (1/9). As the M2 segment lines the channel pore, it is reasonable to hypothesize that variants in the M2 will lead to significant perturbation of the channel structure, affecting the passage of chloride through the pore. Similarly, individuals with variants in the M1 and M3 mainly had focal seizure types with early onset and severe ID, confirming the crucial function of these domains.

### Treatment implications

Treatment was only effective in a limited number of individuals. These were treated according to their seizure types, as currently there is no precision medicine available for GABA_A_Rs. Even if benzodiazepines target the receptor, the sedative effects of these medications render them less useful for long-term treatment. Additionally, it was recently shown that VGB should be avoided in certain individuals as it can aggravate symptoms^22^. Seizure freedom was more common in individuals with variants in the ECD or truncating variants, most likely because of their milder phenotype. However, only 36% and 40%, respectively, achieved seizure freedom, highlighting the severity of disease and limited treatment options.

A novel drug targeting the GABA_A_R is showing promising results in individuals with *PCDH19*-related epilepsy (ClinicalTrials.gov Identifier: NCT02358538). The neurosteroid ganaxolone is a positive allosteric modulator of the GABA_A_R and as such it would theoretically be a relevant ASM in individuals with *GABRB3* variants. Future studies are needed to investigate these theories further.

### Perspectives

It is widely accepted that pathogenic variants in the GABA-related genes cause loss-of-function of the resulting receptors. This leads to disinhibition in the GABAergic neurons, and thus increased neuronal excitability. Numerous studies have shown that variants in both the ECD and TMD cause loss-of-function and lead to severe phenotypes in individuals^3, 32^. In contrast to this, two variants in *GABRB3* were recently shown to lead to gain-of-function receptors^22^. Interestingly, the affected individuals showed hypersensitivity to VGB, a GABA enhancer, adding treatment response to the list of implications associated with functional effects of *GABRB3* variants. In this study, we observed clear differences in the phenotypic severity depending on the structural location of the variants. This suggests that the functional effects of variants in for example the TMD differs from those in the ECD, and that equating the impaired function caused by missense variants to the loss of function caused by protein truncating variants is an oversimplification. More patients are expected to be found and aggregated as genetic test are expanding. Considering that the estimated incidence for pathogenic de novo variants within *GABRB3* is 2.1 per 100.000 live births, we can expect just in the US at least 81.9 new cases per year^33^. Future functional characterization of variants is needed to investigate the hypothesis presented here, and to determine whether the associations between genetic location and phenotype found here also correlates with functional effects. Such studies could potentially also elucidate the relationship between variant location, phenotype and treatment response paving the way for a personalized medicine approach.

## Data Availability

De-identified data will be made available to those eligible. Data includes the GABRB3 database and data used for 3D analysis including statistical logging. Data will be stored for six months.

## Data availability

De-identified data will be made available to those eligible per request to the corresponding authors.

## Acknowledgements

We thank the individuals and their families for participating in this study.

AM and MA are partly funded by the NIHR GOSH BRC. AM is funded by MRC and the Rosetrees Trust. The views expressed are those of the author(s) and not necessarily those of the NHS, the NIHR or the Department of Health. The Cambridge NIHR Bioresource provided support in this study. The DDD study presents independent research commissioned by the Health Innovation Challenge Fund [grant number HICF-1009-003], a parallel funding partnership between Wellcome and the Department of Health, and the Wellcome Sanger Institute [grant number WT098051]. The views expressed in this publication are those of the author(s) and not necessarily those of Wellcome or the Department of Health. The study has UK Research Ethics Committee approval (10/H0305/83, granted by the Cambridge South REC, and GEN/284/12 granted by the Republic of Ireland REC). The research team acknowledges the support of the National Institute for Health Research, through the Comprehensive Clinical Research Network.

## Author information

Conceptualization: KMJ, SI, MG, PS, RS, EG

Data curation: KMJ, SI, MG, NAM, EP, ES, ADSM, MTA, AM, RP, AP, MK, GA, FH, ASJ, MD, KJ, LDW, EHB, NEV, MvK, WF, GV, PS, FZ, HMHB, MH, ME, DS, UV, TS, JRL, KP, JK, KMK, PYBA,PS, KS, JK, MT, MD, AD, DLa, DLe, AJC, VWY, PKA, RS, EG

Formal analysis: KMJ, SI, MG, EG

Funding acquisition: PKA, RS

Investigation: KMJ, SI, MG, EG

Methodology: KMJ, SI, MG, NAM, AJC, VWY, PKA, RS, EG

Project administration: Not relevant

Resources: Not relevant

Software: Not relevant

Supervision: DLa, PS, AJC, PKA, RS, EG

Validation: Not relevant

Visualization: KMJ, SI

Writing – original draft: KMJ, SI, MG, EG

Writing – review & editing: KMJ, SI, MG, NAM, EP, ES, ADSM, MTA, AM, RP, AP, MK, MD, KJ, LDW, EHB, NEV, MvK, WF, GV, HMHB, PS, FZ, MH, ME, DS, UV, TS, JRL, KP, JK, KMK, PYBA, PS, KS, JK, MT, MD, AD, DLa, DLe, AJC, VWY, PKA, RS, EG

## Ethics declaration

All institutions involved in human participant research received local IRB approval (main IRB: The ethics committee of Region Zealand, Denmark). Written informed consent, including authorization for reproduction of video images, was obtained for all individuals (or legal guardians) and family members where necessary. Individual data were collected according to local ethics committee guidelines.

## Supplementary material

Table E1: Phenotypes of individuals with pathogenic *GABRB3* variants.

